# Perceived Healthcare Quality, Patient Satisfaction, and Loyalty in Egypt’s Health Insurance Organization: A Multivariate Analysis

**DOI:** 10.64898/2026.01.05.26343430

**Authors:** Doaa S. Galal, Magda R. Ahmed, Ekram W. Abd El-Wahab, Basem F. Abdel-Aziz

## Abstract

**Background:** Patient satisfaction and loyalty are key indicators of healthcare quality and are influenced by multiple service domains. Understanding which aspects of care drive these outcomes is essential for Health Insurance Organization (HIO) hospitals aiming to improve patient experience.

**Methods:** An analytical cross-sectional study was conducted across four hospitals in Alexandria, Egypt. A total of 983 patients were recruited using stratified random sampling from various hospital departments. Data was collected through structured face-to-face interviews using a validated Arabic National Patient Satisfaction Survey covering eight healthcare quality domains. Multivariate linear regression models were fitted to assess the impact of these domains on patient satisfaction and loyalty. Mediation analysis examined the indirect effect of satisfaction on the relationship between quality and loyalty.

**Results:** The regression model significantly predicted patient satisfaction (Adjusted R² = 0.573, F(8, 974) = 165.93, p < 0.001), with Information, Empathy, and Organization emerging as the strongest predictors. All eight service quality dimensions contributed positively, and no multicollinearity issues were detected. The hierarchical model predicting loyalty was significant, explaining 52.5% of the variance in Model 1 and increasing to 64.8% after adding satisfaction (F(9, 973) = 198.59, p < 0.001). Patient satisfaction became the dominant predictor of loyalty, and the effect of Transition became non-significant once satisfaction was included, indicating full mediation. Overall, satisfaction mediated the relationship between service quality and loyalty, substantially strengthening predictive power.

**Conclusion:** Effective communication, clinical competency, organizational consistency, and responsive care are central drivers of patient satisfaction and loyalty in HIO hospitals. Enhancing these domains may improve patient retention and overall healthcare experiences.

**Study highlights:**

- Provides actionable priorities for clinicians and managers, showing that improving communication, empathy, and care organization yields the largest gains in patient satisfaction.
- Identifies system-level gaps (waiting times, discharge transition, payment processes) that hospital managers and policymakers can directly target for quality improvement.
- Demonstrates that patient satisfaction is the main driver of loyalty, informing policy decisions on where investments will have the greatest impact.
- Uses a large, multicenter sample and validated tool, offering reliable, generalizable evidence to guide HIO performance monitoring and reform.

## INTRODUCTION

Healthcare quality is widely acknowledged as a cornerstone of effective health systems and a major determinant of clinical outcomes, patient experience, and overall system sustainability. International organizations such as the World Health Organization (WHO), the Institute of Medicine (IOM), and the Agency for Healthcare Research and Quality (AHRQ) define high-quality healthcare as care that is safe, effective, patient-centered, timely, efficient, and equitable (1-3). These multidimensional attributes highlight the complexity of healthcare delivery and underscore the importance of patient perceptions in evaluating service performance. As the direct recipients of care, patients are uniquely positioned to assess interpersonal communication, service accessibility, care coordination, and environmental aspects of healthcare facilities, making their evaluations critical to comprehensive quality assessment (4).

Perceived healthcare quality has repeatedly been shown to influence patient satisfaction, which represents the degree to which healthcare services meet or exceed patients’ expectations (5). Satisfaction is not only a reflection of experiences but is also linked to behavioral and clinical outcomes, such as treatment adherence, continuity of care, utilization patterns, and health improvement. Consequently, patient satisfaction has become an essential patient-reported outcome measure in health system performance assessment. In parallel, patient loyalty, which encompasses intentions to revisit the healthcare provider and recommend services to others, emerges as a key indicator of long-term organizational sustainability. Improvements in loyalty can strengthen system trust, enhance service uptake, and promote stable demand, particularly within public insurance systems where retention is vital (6).

Empirical studies have demonstrated robust associations among perceived healthcare quality, patient satisfaction, and loyalty. Healthcare quality often exerts both direct effects on loyalty and indirect effects mediated through satisfaction, making satisfaction a central pathway in the quality–loyalty relationship (7). Evidence from Middle Eastern and developing health systems consistently supports this model, showing that improvements in service quality dimensions, such as responsiveness, communication, and provider competence, significantly enhance satisfaction and, subsequently, loyalty (8).

In Egypt, the Health Insurance Organization (HIO) plays a critical role in delivering services to a large segment of the population. Despite recent reforms aimed at expanding coverage, upgrading infrastructure, and improving service delivery, several studies have highlighted persistent gaps in perceived quality and satisfaction among HIO beneficiaries (9). Issues related to accessibility, provider competence, waiting times, communication, facility conditions, and accessibility remain recurrent concerns that may influence beneficiaries’ satisfaction and continued utilization. As Egypt advances toward universal health insurance, understanding the mechanisms linking perceived quality, satisfaction, and loyalty becomes increasingly important for improving service performance and policy planning. Recent studies in Egypt indicate that improvements in facility conditions, provider competence, communication, and accessibility enhance satisfaction and loyalty among Health Insurance Organization beneficiaries (10, 11). Understanding these relationships is crucial for guiding reforms under Egypt’s Universal Health Insurance system.

Against this backdrop, the present study examines how multiple domains of perceived healthcare quality influence patient satisfaction and loyalty within HIO hospitals in Alexandria. Using multivariate analytical techniques, the study aims to clarify these interrelationships and contribute evidence to guide ongoing improvements within Egypt’s health insurance system.

## METHODS

### Study Design, Setting, and Population

The study was conducted from June 2022 to December 2023 across four HIO hospitals in Alexandria. Data was collected through patient interviews in inpatient wards, outpatient clinics, emergency departments, and other service units. An analytical cross-sectional design was employed, targeting patients attending these hospitals from September 2022 to December 2023. Eligible participants included HI beneficiaries and parents of pediatrics patients at, while those declining participation were excluded.

### Sampling Design and Technique

Simple stratified random sampling was applied, with hospitals serving as strata. Participants were selected from inpatient, outpatient, emergency, and specialized services (e.g., renal dialysis). Equal distribution across hospitals was maintained until the required sample was obtained. A power analysis for multiple regression was performed using G*Power (α = 0.05, power = 0.80) for the overall F-test (linear multiple regression: R² deviation from zero) with 87 predictors. Based on conventional Cohen effect sizes, the estimated required sample sizes were approximately 481 for a small effect (f² = 0.02), 141 for a medium effect (f² = 0.15), and 111 for a large effect (f² = 0.35). To ensure adequate power for detecting even small effects and to accommodate potential attrition, the final target sample size was set at 900 participants.

### Data Collection Methods and Tools

Face-to-face interviews were conducted using a structured questionnaire (File S1). Section one captured sociodemographic characteristics (age, sex, marital and work status, residence, family size, insurance coverage, occupation, education, income), current acute or chronic health conditions, and health insurance utilization patterns (hospital, department, service duration, visit frequency). Section two consisted of the validated Arabic National Patient Satisfaction Survey (NPSS), a modified SERVQUAL tool used in previous UAE studies (12, 13). The NPSS includes eighty-seven items covering healthcare quality, patient satisfaction, and patient loyalty, with reported Cronbach’s alpha of 0.961 (14). Items were rated on a five-point Likert scale (1 = strongly disagree to 5 = strongly agree).

A pilot test with fifteen participants confirmed clarity of wording. Cronbach’s alpha demonstrated strong internal consistency: healthcare quality (0.924), patient satisfaction (0.821), and patient loyalty (0.731).

### Assessment of Healthcare Service Quality

Healthcare service quality was assessed using eighty-seven items distributed across eight domains encompassing key aspects of patient experience. These domains included tangibles and facilities, which evaluated physical conditions and ancillary services; courtesy and empathy, addressing interpersonal interactions; and responsiveness along with physiological and psychosocial aspects, capturing responsiveness and patient involvement. Additional domains assessed competency, fairness, and trust in providers; information and communication; and availability, accessibility, and timeliness, which covered waiting times, delays, and service access. The evaluation also included patients’ transition to home following discharge and organization management, regulation, and payment, reflecting broader administrative and financial processes. Mediation analysis was performed using regression-based approaches to explore indirect effects.

### Data Management and Screening

Data was entered and managed in SPSS (version 29). Questionnaires with more than 10% missing items (17 of 1,000) were excluded. Remaining missing data were inspected, and imputation methods were applied based on the extent and mechanism of missingness: mean imputation for items with minimal missingness and multiple imputation for variables with nontrivial missingness. Continuous variables were assessed for normality using skewness and kurtosis statistics as well as visual inspections. Likert-scale items were treated as continuous for multivariate analyses after confirming acceptable distributional properties.

### Scale Validation

Construct reliability and dimensionality were assessed using Cronbach’s alpha and exploratory factor analysis (EFA). Principal-axis factoring with oblique rotation was used to explore the latent structure of the healthcare-quality instrument. Factors were retained based on eigenvalues greater than 1, scree plots, and interpretability. Items with low loadings (<0.40) or cross-loadings were considered for removal. Scale scores were computed as the mean of retained items per domain (range 1–5) and interpreted using thresholds adapted from Wu (2007): very low (1.00–1.79), low (1.80–2.59), moderate (2.60–3.39), good (3.40–4.19), and very good (4.20–5.00).

### Descriptive Analysis

Participant characteristics and questionnaire responses were summarized using frequencies and percentages for categorical variables and means ± standard deviations for continuous variables. Item-level responses were tabulated as counts and percentages.

### Multivariate Analyses

Associations between healthcare-quality domains and outcomes (patient satisfaction and loyalty) were examined using multiple linear regression for continuous outcomes, multivariable logistic regression for binary outcomes, and multivariate analysis of variance (MANOVA) for joint testing of correlated dependent variables. Models included relevant sociodemographic covariates (age, sex, education, income, insurance usage) and hospital fixed effects to account for clustering. Variable selection was guided by theoretical relevance and univariable screening (p < 0.20). Interaction terms were included as indicated by theory, and model fit was evaluated using adjusted R² (linear models) or pseudo-R² (logistic models).

### Model Diagnostics and Assumptions

Model assumptions were assessed, including linearity, independence of errors, homoscedasticity, normality of residuals, and multicollinearity (variance inflation factor, VIF). For violations, robust standard errors or alternative estimation methods (e.g., generalized estimating equations) were employed. Statistical significance was set at α = 0.05, with 95% confidence intervals reported.

## RESULTS

### Characteristics of the study population

Participants were recruited from a range of hospital units, primarily from inpatient departments (48.9%), followed by outpatient clinics (34.5%) and emergency departments (13.1%), with only a small number drawn from other services such as dialysis and physiotherapy units (Table S1).

The sample comprised 57.6% females and 42.4% males, with the participants largely urban residents (73.1%). The mean age was 45.1 ± 11.05 years, with the majority between in the age group 30–50 years. Over half of the respondents were employed and reported inadequate income. Educational attainment was generally low to intermediate, with most participants reporting illiteracy or intermediate schooling and only 11.9% having university-level education. Most participants were married (67.2%), lived in households of three to four members, and had 1–4 dependents enrolled in the HIO.

Chronic and acute health problems were reported at comparable levels, with 54.6% of participants identifying chronic conditions and 45.4% reporting acute issues. The majority had been using HIO services for more than two years, reflecting a stable beneficiary population. Frequent service utilization was common, with nearly 60% having used services up to three times, while approximately 40% reported four or more visits (Table 1).

**Table 1:** Characteristics of the study population.

| Characteristics | n | % |
| --- | --- | --- |
| Sex |  |  |
| Male | 423 | 42.3 |
| Female | 577 | 57.7 |
| Age (Years) |  |  |
| <20 | 4 | 0.4 |
| 20–<40 | 360 | 36.6 |
| 40–60+ | 619 | 63.0 |
| Mean±SD, Range | 45.1±11.05, 20-70 |  |
| Residency |  |  |
| Rural | 264 | 26.9 |
| Urban | 719 | 73.1 |
| Employment |  |  |
| Yes | 515 | 52.4 |
| No | 468 | 47.6 |
| Education Level |  |  |
| Illiterate/read and write | 234 | 23.8 |
| Intermediate (Primary/Secondary) | 632 | 64.3 |
| High (University) | 117 | 11.9 |
| Income |  |  |
| Adequate (Enough / Saving) | 403 | 41 |
| Inadequate (Not Enough / Loans) | 945 | 59 |
| Marital Status |  |  |
| Married | 661 | 67.2 |
| Not married (Single / Widow / Divorced) | 322 | 32.8 |
| Family Size |  |  |
| 1–4 | 676 | 68.8 |
| 5+ | 307 | 31.2 |
| Dependents Covered by Insurance |  |  |
| 1–4 dependents | 924 | 92.4 |
| ≥5 dependents | 76 | 7.6 |
| Health problem |  |  |
| Chronic | 537 | 54.60 |
| Acute | 446 | 45.40 |
| Duration of health insurance service utilization |  |  |
| 1–2 years | 91 | 9.30 |
| 2–6+ | 909 | 90.70 |
| Frequency of service utilization |  |  |
| ≤3 visits | 588 | 59.80 |
| ≥4 visits | 395 | 40.20 |

SD; Standard Deviation

### Patient Perceptions of Healthcare Quality

The perceptions of healthcare quality varied across the domains assessed. Tangible aspects such as cleanliness, staff appearance, and facility adequacy, received moderate agreement, with endorsement levels generally ranging from 40% to 46%. Ancillary services such as parking, communication facilities, nutritional services, and waiting areas received similarly moderate ratings. Empathy and courtesy indicators showed comparable patterns, with roughly 45% of patients perceiving respectful interactions and privacy maintenance. Responsiveness and psychological support were weaker, with fewer than half reporting adequate emotional support or timely engagement from providers.

Patient involvement in care decisions, clarity of explanations, and justification for delays showed moderate ratings but remained below 50%. Fairness, trust, and consistency of information were also suboptimal, with fewer than half perceiving equitable treatment or reliable communication from staff. Confidence in provider competency was moderate, with 43–45% expressing trust in doctors, nurses, and administrative staff. Communication and information provision were similarly rated, with 42–46% agreeing that they received sufficient details regarding their condition and treatment. Timeliness and waiting times were key concerns, as less than half reported timely procedures, test results, or discharge processes. Availability of physicians and accessibility of services were moderate overall but lower during weekends and for specialized care. Transition to home was notably weak, with only one-quarter to one-third receiving adequate discharge or recovery instructions. Management-related domains were mixed, with unnecessary tests, strict rules, and coordination issues frequently reported. Payment-related experiences were among the lowest-rated domains, particularly regarding simplicity and perceived fairness of financial procedures.

Approximately 42–46% of patients expressed satisfaction with treatment outcomes and overall services, while loyalty indicators, such as willingness to return or recommend the hospital, were slightly lower, with nearly one-third expressing reluctance to recommend HIO facilities (Table S2).

### Evaluation of Service Quality Domains

Domain-level service quality scores indicated moderate satisfaction overall, with tangibles, empathy, and organizational management achieving the highest positive ratings (Table 2). Most patients (61.7%) rated overall service quality as moderate, and 42% reported moderate-to-high satisfaction. Loyalty was also predominantly moderate or good, though 24.9% of participants reported low loyalty (Table 3).

**Table 2:**
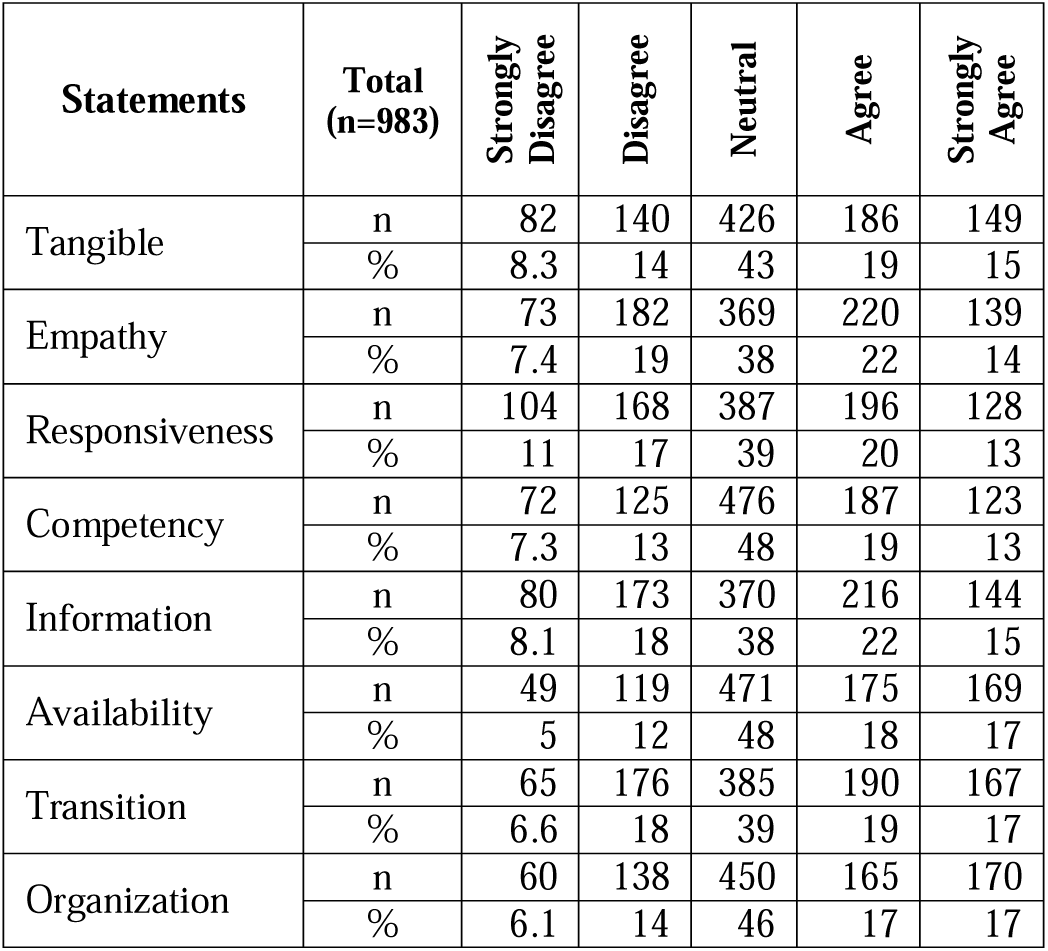
Evaluation of Health Care Service Quality Domains.

**Table 3:** Overall Evaluation of Health Care Service Quality.

| Statements |  | Strongly Disagree | Disagree | Neutral | Agree | Strongly Agree |
| --- | --- | --- | --- | --- | --- | --- |
| Overall quality evaluation | n | 13 | 96 | 607 | 198 | 69 |
|  | % | 1.3 | 9.8 | 61.7 | 20.1 | 7.0 |
| Statements |  | Very Unsatisfied | Unsatisfied | Moderately satisfied | Satisfied | Very satisfied |
| Patient Satisfaction | n | 72 | 162 | 337 | 215 | 197 |
|  | % | 7.3 | 16.5 | 34.3 | 21.9 | 20.0 |
| Statements |  | Very low | Low | Moderate | Good | Very good |
| Patient Loyalty | n | 105 | 140 | 344 | 173 | 221 |
|  | % | 10.7 | 14.2 | 35.0 | 17.6 | 22.5 |

### Reliability of Healthcare Quality Constructs

All quality, satisfaction, and loyalty constructs demonstrated strong internal consistency (Cronbach’s α > 0.80). The highest reliability was observed for the overall healthcare quality construct (α = 0.937), whereas patient loyalty showed acceptable reliability (α = 0.769). Information and communication and availability of resources were among the most positively rated domains based on mean scores (Table S3).

### Multivariate Analysis for Predicting Patient Satisfaction

A multiple linear regression analysis was conducted to identify predictors of patient satisfaction. The overall model was statistically significant, F(8, 974) = 165.93, p < .001, explaining 57.7% of the variance in satisfaction (Adjusted R² = 0.573), indicating strong predictive capacity (Table 4).

**Table 4:** ANOVA results for predictors of patient satisfaction.

| **Source** | **Sum of Squares** | **df** | **Mean Square** | **F** | **Sig.** |
| --- | --- | --- | --- | --- | --- |
| Regression | 643.167 | 8 | 80.396 | 165.927 | < .001 |
| Residual | 471.929 | 974 | 0.485 |  |  |
| Total | 1115.097 | 982 |  |  |  |

All eight service quality dimensions, Tangibility, Empathy, Competence, Involvement, Organization, Availability, Information, and Transition, were significant positive predictors of patient satisfaction. Among these, Information (β = 0.326), Empathy (β = 0.233), and Organization (β = 0.161) emerged as the strongest contributors to the model. Collinearity diagnostics confirmed minimal multicollinearity (VIF range: 1.071–1.392). The Durbin–Watson statistic of 1.028 indicated some positive autocorrelation but remained within an acceptable range for large samples (Table 5 and Table S4).

**Table 5:** Regression Coefficients for predictors of patient satisfaction.

| **Predictor** | **B (Unstd.)** | **SE** | **Beta (Std.)** | **t** | **Sig.** | **Tolerance** | **VIF** |
| --- | --- | --- | --- | --- | --- | --- | --- |
| Constant | -1.340 | 0.141 | — | -9.508 | < .001 | — | — |
| Tangibility | 0.146 | 0.025 | 0.130 | 5.856 | < .001 | 0.877 | 1.141 |
| Empathy | 0.251 | 0.026 | 0.233 | 9.487 | < .001 | 0.719 | 1.392 |
| Competence | 0.163 | 0.025 | 0.146 | 6.412 | < .001 | 0.838 | 1.193 |
| Involvement | 0.125 | 0.022 | 0.124 | 5.618 | < .001 | 0.892 | 1.121 |
| Organization | 0.178 | 0.024 | 0.161 | 7.456 | < .001 | 0.933 | 1.071 |
| Availability | 0.122 | 0.025 | 0.109 | 4.860 | < .001 | 0.865 | 1.156 |
| Information | 0.343 | 0.025 | 0.326 | 13.649 | < .001 | 0.763 | 1.310 |
| Transition | 0.104 | 0.026 | 0.098 | 4.012 | < .001 | 0.736 | 1.359 |

Overall, the results highlight that improvements in communication, empathy, organizational flow, and staff competence have the greatest impact on enhancing patient satisfaction within the studied hospitals. The collinearity diagnostics are provided in Table S5. Examination of the residuals showed no violations of regression assumptions, with no evidence of heteroscedasticity.

### Prediction of Loyalty and Mediation Effect of Patient Satisfaction

A hierarchical regression analysis was performed to examine the predictors of patient loyalty and to test the mediating role of patient satisfaction (Table 6).

**Table 6.** ANOVA results for predictors of patient loyalty.

| **Model** | **Source** | **Sum of Squares** | **df** | **Mean Square** | **F** | **Sig.** |
| --- | --- | --- | --- | --- | --- | --- |
| Model 1: Service Quality Dimensions → Loyalty | Regression | 674.526 | 8 | 84.316 | 134.710 | < .001 |
|  | Residual | 609.631 | 974 | 0.626 |  |  |
|  | Total | 1284.157 | 982 |  |  |  |
| Model 2: Service Quality Dimensions + Satisfaction → Loyalty | Regression | 831.497 | 9 | 92.389 | 198.591 | < .001 |
|  | Residual | 452.659 | 973 | 0.465 |  |  |
|  | Total | 1284.157 | 982 |  |  |  |

Dependent Variable: Loyalty

Model 1 included the eight service quality dimensions and significantly predicted loyalty, F(8, 974) = 134.71, p < .001, accounting for 52.5% of the variance (Table 6). The most influential predictors were Information (β = .356), Competence (β = .175), Empathy (β = .173), and Organization (β = .170), as detailed in Table 7 and Table S6.

**Table 7.**
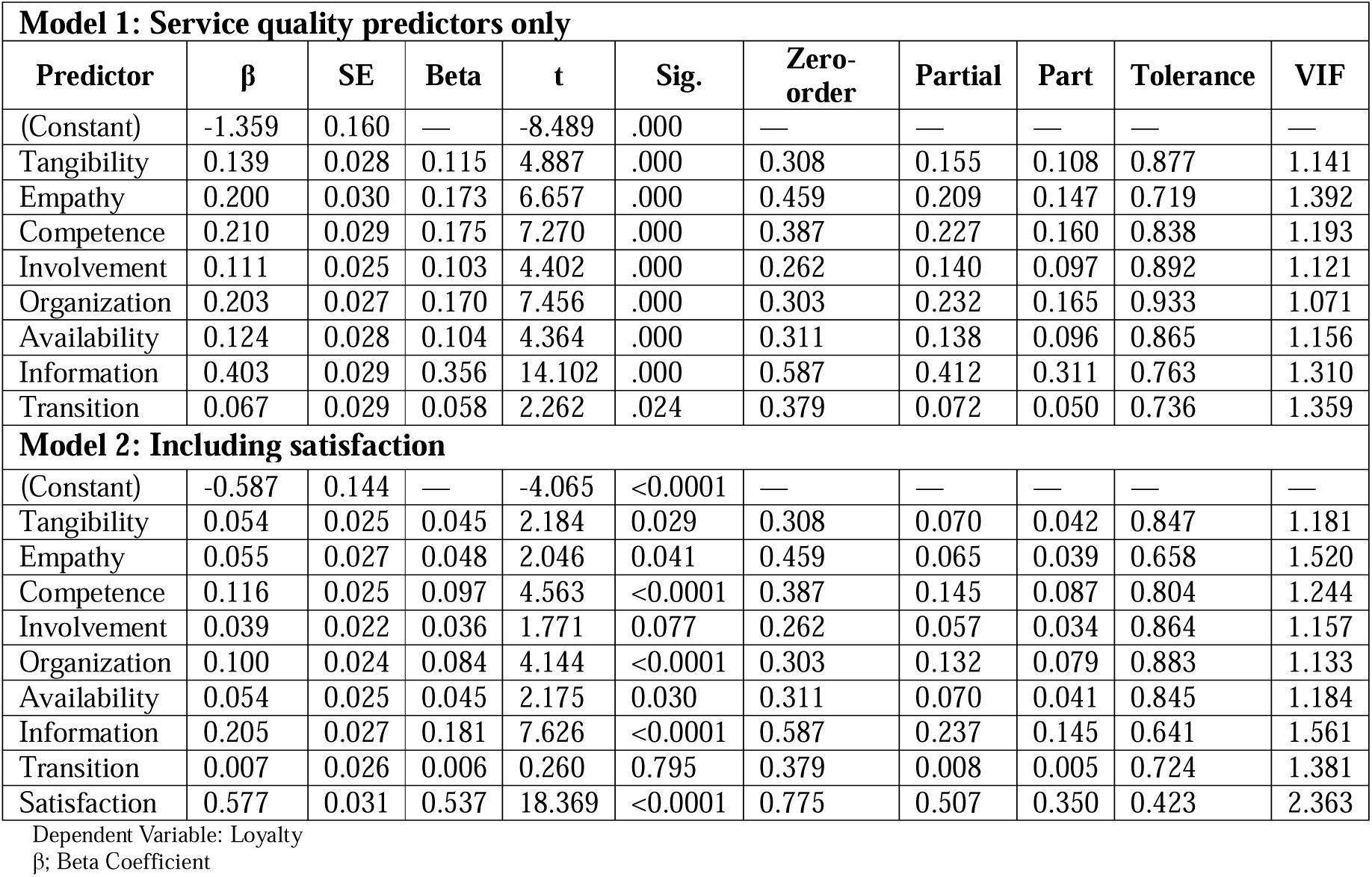
Regression Coefficients for predictors of patient loyalty.

In Model 2, patient satisfaction was added to the predictors. The model’s explanatory power increased substantially to 64.8%, F(9, 973) = 198.59, p < .001. Satisfaction emerged as the strongest predictor of loyalty (β = .537, p < .001). After controlling for satisfaction, the effects of most service quality dimensions were reduced but remained significant. Transition, however, became non-significant (p = .795), indicating a full mediation effect of satisfaction on the relationship between transition quality and loyalty (Table 7 and Table S6).

## DISCUSSION

The findings of this study highlight the pivotal role of perceived healthcare quality in shaping both patient satisfaction and loyalty within hospitals under the HIO. In alignment with a robust body of international literature, our analysis identified key domains as core drivers, in particular information quality, clinical competency, organizational procedures (rules), responsiveness, availability, and transition, as major predictors of satisfaction and loyalty. Moreover, the mediation analysis highlights satisfaction as a crucial mechanism transferring the effect of perceived quality to loyalty, underscoring its role as a mediator rather than simply a parallel outcome. Such insights are highly relevant for health insurance hospitals aiming to improve patient experience and retention under Egypt’s ongoing reforms (15, 16).

The most consistent and powerful predictor in our models was the quality of information provided to patients. This domain had the strongest effect on both patient satisfaction and patient loyalty. Indeed, accurate, clear, and timely information about care and procedures is fundamental to patients’ trust and confidence in their healthcare providers. When hospitals communicate well, patients are more likely to feel understood, valued, and involved in their care. This finding accords with prior studies in Egypt and Saudi Arabia showing that adequacy of information is a strong determinant of satisfaction and loyalty (17, 18).

Closely following information quality were the domains of competency (clinical skill and staff performance) and organizational rule adherence (consistency, fairness, and clarity of hospital procedures). Their relative prominence in predicting satisfaction and loyalty suggests that patients value not only good communication but also reliable, skilful, and fair service delivery. The literature on hospital service quality consistently highlights assurance (competence) and reliability (rule consistency) as central to building trust, satisfaction, and loyalty across diverse healthcare settings (19, 20). In our context–a public insurance hospital network serving a broad socio-economic population–ensuring consistent, competent care and transparent procedures may be especially important to retain patient trust and allegiance.

Our mediation analysis confirms a widely proposed model in quality–satisfaction–loyalty research. Perceived quality significantly influences satisfaction, which in turn drives loyalty intentions. The indirect effect via satisfaction accounted for nearly half of the total effect of quality on loyalty. In other words, much of quality’s influence on loyalty works by shaping how satisfied patients feel about their care, rather than directly affecting loyalty in isolation. This pattern echoes findings from several empirical studies in different contexts: a multicenter study from China demonstrated that service quality affects loyalty primarily through satisfaction, with both direct and mediated effects present but the mediated path stronger (20). Similarly, studies in hospital contexts in Southeast Asia have reported that patient satisfaction mediates between service quality (or its components) and loyalty or revisit intentions (15, 16). This mediated relationship has practical implications. For instance, efforts to improve perceived quality should aim not only at objective improvements (facilities, staffing, procedures) but also at optimizing patient-perceived experiences, especially around communication, trust-building, and service reliability, which drive satisfaction and, ultimately loyalty. Simply improving physical infrastructure or technical capacity might not yield sustained loyalty if communication and service consistency remain poor.

Interestingly, the tangible and empathy domains, despite their significant contribution to the model, showed comparatively modest effects in predicting satisfaction and loyalty. Better infrastructure, modern equipment, or more welcoming physical environments had a weaker influence than informational and competence-related domains. Similarly, expressions of courtesy and empathy, though positively perceived by almost half of respondents, did not drive satisfaction or loyalty as strongly as elements tied to communication, reliability, and competence.

This does not imply that environment or empathy are less important. Other research, including in Egypt, finds that facility conditions and staff behavior remain important for patient satisfaction and can influence perceived value of care, especially when other domains are weak (18, 21). However, our results suggest that in the context of HIO hospitals, which are often resource-constrained and serving high patient loads, beneficiaries may prioritize clarity, competence, and consistency over comfort or interpersonal warmth.

These findings have a particular relevance in the context of Egypt’s expanding health insurance coverage. Past studies in Egyptian public hospitals have reported adequate overall satisfaction when service quality was assessed via modified SERVQUAL instruments; yet facility environment and service procedures frequently ranked among the lowest scored domain (18, 21). Our study reinforces this evidence showing that even when perceived quality is moderate, improvement in information, organizational processes, and clinical competency can yield meaningful gains in satisfaction and loyalty. Given that many beneficiaries of HIO are from lower-income or underserved populations, ensuring clarity about procedures, reliable service delivery, and transparent communication may foster equity, trust, and sustained engagement with the insurance system.

Moreover, as the system scales up, high patient volumes and resource pressure may limit investments in infrastructure or aesthetics. Our data suggests that strategic investment in training staff to communicate effectively, standardizing procedures, and strengthening clinical competence which are potentially more cost effective than extensive physical upgrades, could deliver more value in terms of patient retention and satisfaction. This is consistent with recent recommendations from quality improvement frameworks for public hospitals in post-reform settings (19).

The findings in this study provide key guidance and actionable insights for clinical practice and policy development. Health insurance hospital managers and policymakers should focus on interventions that enhance communication, clinical competency, and organizational consistency to improve patient satisfaction and loyalty. Efforts should ensure that patients receive clear, timely, and accurate information regarding diagnoses, treatment plans, service procedures, and discharge instructions, facilitated through staff training, standardized information materials, and patient navigators (17, 18). Continuous professional development for clinical and administrative staff is essential. Equally important is the standardization and fair enforcement of organizational procedures, including admission, discharge, payment, and service prioritization, to foster trust. Optimizing patient flow and responsiveness by minimizing waiting times, ensuring timely service delivery, and addressing patient concerns or psychosocial needs can further enhance satisfaction and loyalty, even where infrastructure improvements are limited (16, 19). Regular monitoring through satisfaction and loyalty surveys can help identify gaps and guide ongoing quality improvement initiatives.

In conclusion, perceived healthcare quality, especially in domains of information provision, clinical competency, responsiveness, availability, and organizational consistency, significantly influences patient satisfaction and loyalty in HIO hospitals. Physical environment and empathy contribute modestly. The mediation effect of satisfaction underscores its role as an intermediary mechanism. These findings support targeted quality-improvement strategies emphasizing communication, staff competence, and procedural transparency as efficient, impactful levers for improving patient experience and loyalty, vital for the sustainability of Egypt’s health insurance hospitals (15, 21).

### Limitations and Strengths

Despite its strengths, including a large hospital-based sample and comprehensive assessment across multiple quality domains, this study has limitations. First, the cross-sectional design prevents inference of causality; while statistical mediation suggests directional relationships, longitudinal studies would better confirm the dynamic links between quality perceptions, satisfaction, and loyalty over time. Second, the reliance on patient self-report introduces potential biases; e.g., social desirability, recall bias, or response bias, which may inflate satisfaction or loyalty ratings. Third, generalizability may be limited to the HIO hospital context in Alexandria. Private hospitals or other governorates might yield different patterns. Finally, while the questionnaire demonstrated high internal consistency, some domains (e.g., transition to home) may be sensitive to the timing of discharge or follow-up, potentially affecting the validity of those subscales.

Nevertheless, key strengths of this study include its comprehensive domain assessment, robust multivariate modelling, and large sample size, enabling detection of relatively modest but meaningful predictors. The mediation analysis adds value by clarifying the mechanism through which perceived quality translates into loyalty, an insight often assumed but not always empirically tested.

## Data Availability

All data produced in the present study are available upon reasonable request to the authors

## Ethical consideration

### Ethical approval and consent to participate

The study was approved by the institutional review board and the ethics committee of the High Institute of Public Health affiliated with Alexandria University, Egypt. We sought the permission and support of the local health authorities to conduct the study in the selected districts in Alexandria. The study was conducted in accordance with the international ethical guidelines and of the Declaration of Helsinki. Informed written consent was obtained from each participant after explaining the aim and concerns of the study. Data sheets were coded by number to ensure anonymity and confidentiality of the participants’ data.

This article does not contain any studies with animals performed by any of the authors.

- **Informed consent:** Declared
- **Funding:** None
- **Conflicts of interest:** None to declare.

### Consent for publication

All authors approved the manuscript for publication

### Availability of supporting data

All data are fully available without restriction by the corresponding author at

## Acknowledgements

We would like to acknowledge the study participants for accepting to participate in the study.

## Notes

### Competing Interest Statement

The authors have declared no competing interest.

### Funding Statement

This study did not receive any funding

### Author Declarations

The study was approved by the institutional review board and the ethics committee of the High Institute of Public Health affiliated with Alexandria University, Egypt. We sought the permission and support of the local health authorities to conduct the study in the selected districts in Alexandria. The study was conducted in accordance with the international ethical guidelines and of the Declaration of Helsinki. Informed written consent was obtained from each participant after explaining the aim and concerns of the study. Data sheets were coded by number to ensure anonymity and confidentiality of the participant data. This article does not contain any studies with animals performed by any of the authors.

