## Supplementary material for "Perceived Healthcare Quality, Patient Satisfaction, and Loyalty in Egypt’s Health Insurance Organization: A Multivariate Analysis": Suppl tables

Table S1: Distribution of the study sample per department

| **Department Name** | **Total participants (n=**983) | |
| --- | --- | --- |
|  | **n** | **%** |
| Inpatient | 481 | 48.9 |
| Outpatient | 339 | 34.5 |
| Emergency department | 129 | 13.1 |
| Other services | 34 | 3.5 |

**Table S2: Patients' Perception of the Quality dimensions**

| Statements | Total (Nn=983) | Strongly Disagree | Disagree | Neutral | Agree | Strongly Agree |
| --- | --- | --- | --- | --- | --- | --- |
| **Patients' Perception of the Tangibles and Physical Attributes** | | | | | | |
| T1. I felt cleanliness throughout the hospital | n | 176 | 134 | 226 | 239 | 208 |
|  | % | 17.9 | 13.6 | 23.0 | 24.3 | 21.2 |
| T2. The cabins/wards were clean | n | 166 | 162 | 256 | 221 | 178 |
|  | % | 16.9 | 16.5 | 26.0 | 22.5 | 18.1 |
| T3. The doctors, nurses, and staff were neat and appeared (dressed professionally) | n | 176 | 145 | 212 | 186 | 264 |
|  | % | 17.9 | 14.8 | 21.6 | 18.9 | 26.9 |
| T4. The hospital had a comfortable rest area for patients | n | 174 | 157 | 204 | 285 | 163 |
|  | % | 17.7 | 16.0 | 20.8 | 29.0 | 16.6 |
| T5. The hospital has modern-looking equipment | n | 166 | 164 | 209 | 273 | 171 |
|  | % | 16.9 | 16.7 | 21.3 | 27.8 | 17.4 |
| T6. The hospital used state-of-the-art medical equipment | n | 164 | 143 | 253 | 225 | 198 |
|  | % | 16.7 | 14.6 | 25.7 | 22.9 | 20.1 |
| T7. The materials associated with the services (towels, etc.) were visually appealing | n | 156 | 127 | 244 | 259 | 197 |
|  | % | 15.9 | 12.9 | 24.8 | 26.3 | 20.0 |
| T8. The physical facilities at the hospital were visually appealing | n | 178 | 172 | 236 | 225 | 172 |
|  | % | 18.1 | 17.5 | 24.0 | 22.9 | 17.5 |
| T9. The toilets were clean | n | 163 | 165 | 256 | 236 | 163 |
|  | % | 16.6 | 16.8 | 26.0 | 24.0 | 16.6 |
| **Patients' Perception of the other facilities and services** | | | | | | |
| T10. Sufficient parking for patients | n | 180 | 170 | 227 | 230 | 176 |
|  | % | 18.3 | 17.3 | 23.1 | 23.4 | 17.9 |
| T11. Communication facilities for the patients (telephones, operators, signals) | n | 170 | 165 | 281 | 215 | 152 |
|  | % | 17.3 | 16.8 | 28.6 | 21.9 | 15.5 |
| T12. Good nutritional services | n | 171 | 109 | 237 | 236 | 230 |
|  | % | 17.4 | 11.1 | 24.1 | 24.0 | 23.4 |
| T13. Comfortable beds/ waiting areas for patients’ sitters/ relatives. | n | 192 | 176 | 194 | 227 | 194 |
|  | % | 19.5 | 17.9 | 19.7 | 23.1 | 19.7 |
| **Patient’s perception of Empathy and courtesy** | | | | | | |
| E1. I feel that the staff (doctors, nurses, etc.) showed a sincere interest in my condition | n | 141 | 163 | 225 | 237 | 217 |
|  | % | 14.3 | 16.6 | 22.9 | 24.1 | 22.1 |
| E2. I felt that the treatment of patients changed with time | n | 161 | 172 | 230 | 207 | 213 |
|  | % | 16.4 | 17.5 | 23.4 | 21.1 | 21.7 |
| E3. I was treated with respect and dignity while I was in the hospital | n | 162 | 128 | 235 | 218 | 240 |
|  | % | 16.5 | 13.0 | 23.9 | 22.2 | 24.4 |
| E4. My family and visitors were treated with respect and dignity | n | 150 | 145 | 225 | 283 | 180 |
|  | % | 15.3 | 14.8 | 22.9 | 28.8 | 18.3 |
| E5. The admissions staff were friendly and helpful | n | 178 | 157 | 220 | 224 | 204 |
|  | % | 18.1 | 16.0 | 22.4 | 22.8 | 20.8 |
| E6. The nursing staff maintained and respected my privacy | n | 157 | 166 | 215 | 225 | 220 |
|  | % | 16.0 | 16.9 | 21.9 | 22.9 | 22.4 |
| E7. The person who cleaned my room was friendly and courteous | n | 157 | 165 | 254 | 236 | 171 |
|  | % | 16.0 | 16.8 | 25.8 | 24.0 | 17.4 |
| **Patient’s perceptions of Responsiveness and psychological aspects** | | | | | | |
| R1. When I had some anxiety or fear, the doctor discussed it with me | n | 190 | 162 | 260 | 174 | 197 |
|  | % | 19.3 | 16.5 | 26.4 | 17.7 | 20.0 |
| R2. When I had some anxiety or fear, the doctor discussed it with me | n | 199 | 182 | 179 | 218 | 205 |
|  | % | 20.2 | 18.5 | 18.2 | 22.2 | 20.9 |
| **Patient’s Perceptions of Involvement** | | | | | | |
| R 3. I feel that the hospital involved me in the decision about my care (when applicable) | n | 185 | 183 | 201 | 209 | 205 |
|  | % | 18.8 | 18.6 | 20.4 | 21.3 | 20.9 |
| R4. I had said enough about the medical treatment given to me | n | 197 | 189 | 202 | 216 | 179 |
|  | % | 20.0 | 19.2 | 20.5 | 22.0 | 18.2 |
| R5. Someone explained to me the reasons why I had to wait to go to my room | n | 156 | 148 | 233 | 237 | 209 |
|  | % | 15.9 | 15.1 | 23.7 | 24.1 | 21.3 |
| R6. The nursing staff explained the treatment in terms I could understand | n | 176 | 179 | 199 | 220 | 209 |
|  | % | 17.9 | 18.2 | 20.2 | 22.4 | 21.3 |
| R7. When I had important questions to ask the nurse, I got answers that I could understand | n | 192 | 177 | 176 | 227 | 211 |
|  | % | 19.5 | 18 | 17.9 | 23.1 | 21.5 |
| R8. When I had important questions to ask the doctors, I got answers that I could understand | n | 184 | 151 | 238 | 207 | 203 |
|  | % | 18.7 | 15.4 | 24.2 | 21.1 | 20.7 |
| **Patient’s Perception of Fairness and Trust** | | | | | | |
| C1. I felt that one doctor said something, while another doctor said something quite different | n | 166 | 177 | 207 | 203 | 230 |
|  | % | 16.9 | 18.0 | 21.1 | 20.7 | 23.4 |
| C2. I felt that one nurse said something, while another nurse said something quite different | n | 159 | 133 | 225 | 231 | 235 |
|  | % | 16.2 | 13.5 | 22.9 | 23.5 | 23.9 |
| C3. I felt that some patients enjoyed first-class treatment while many others did not | n | 150 | 147 | 268 | 237 | 181 |
|  | % | 15.3 | 15.0 | 27.3 | 24.1 | 18.4 |
| C4. I felt that the doctors were talking about me as I was not there | n | 161 | 132 | 285 | 224 | 181 |
|  | % | 16.4 | 13.4 | 29.0 | 22.8 | 18.4 |
| C5. I felt that the hospital was discriminating in its treatment of patients | n | 145 | 154 | 258 | 226 | 200 |
|  | % | 14.8 | 15.7 | 26.2 | 23.0 | 20.3 |
| C6. I felt that the nurses were talking about me as it was not there | n | 158 | 163 | 243 | 197 | 222 |
|  | % | 16.1 | 16.6 | 24.7 | 20 | 22.6 |
| **Patient’s Perception of Competency and Confidence** | | | | | | |
| C7. I felt that the doctors who treated me were highly knowledgeable | n | 139 | 159 | 240 | 252 | 193 |
|  | % | 14.1 | 16.2 | 24.4 | 25.6 | 19.6 |
| C8. I had full confidence and trust in the doctor treating me | n | 162 | 153 | 241 | 217 | 210 |
|  | % | 16.5 | 15.6 | 24.5 | 22.1 | 21.4 |
| C9. I had full confidence and trust in the nurse treating me | n | 149 | 160 | 251 | 225 | 198 |
|  | % | 15.2 | 16.3 | 25.5 | 22.9 | 20.1 |
| C10. I think that the hospital did all it could to help control my pain | n | 176 | 165 | 220 | 252 | 170 |
|  | % | 17.9 | 16.8 | 22.4 | 25.6 | 17.3 |
| C11. The technical/administrative / support staff appeared professional and technically competent | n | 161 | 168 | 219 | 241 | 194 |
|  | % | 16.4 | 17.1 | 22.3 | 24.5 | 19.7 |
| **Patient’s Perception of Communication** | | | | | | |
| I1. It was easy for me to find someone on the hospital staff to talk to about my concerns | n | 151 | 180 | 233 | 181 | 238 |
|  | % | 15.4 | 18.3 | 23.7 | 18.4 | 24.2 |
| I2. My family (or someone close to me) had enough opportunity to talk to my doctors | n | 151 | 153 | 251 | 162 | 266 |
|  | % | 15.4 | 15.6 | 25.5 | 16.5 | 27.1 |
| I3. My family (or someone dose to) had enough opportunity to talk to the nurses | n | 177 | 165 | 229 | 195 | 217 |
|  | % | 18.0 | 16.8 | 23.3 | 19.8 | 22.1 |
| **Patient’s Perception of Information** | | | | | | |
| I4. Adequate information about my condition or treatment was given to my family | n | 143 | 125 | 258 | 244 | 213 |
|  | % | 14.5 | 12.7 | 26.2 | 24.8 | 21.7 |
| I5. I feel that the hospital informed me about the medical options | n | 170 | 158 | 256 | 191 | 208 |
|  | % | 17.3 | 16.1 | 26.0 | 19.4 | 21.2 |
| I6. While I was in a certain unit, I got enough information about my medical condition | n | 150 | 184 | 227 | 215 | 207 |
|  | % | 15.3 | 18.7 | 23.1 | 21.9 | 21.1 |
| I7. While I was in certain units, I got enough information about the medical treatment I needed | n | 152 | 162 | 262 | 224 | 183 |
|  | % | 15.5 | 16.5 | 26.7 | 22.8 | 18.6 |
| **Patient’s Perception of Timely Matters** | | | | | | |
| A1. After checking in at the hospital, I had to wait before going to my room | n | 128 | 183 | 247 | 216 | 209 |
|  | % | 13.0 | 18.6 | 25.1 | 22.0 | 21.3 |
| A2. After I requested pain medicine, it usually took a long time before I got it | n | 131 | 166 | 230 | 241 | 215 |
|  | % | 13.3 | 16.9 | 23.4 | 24.5 | 21.9 |
| A3. After I used the call button, it usually took a long time to get the help I needed | n | 161 | 157 | 218 | 224 | 223 |
|  | % | 16.4 | 16.0 | 22.2 | 22.8 | 22.7 |
| A4. I thought that I had to wait an unnecessarily long time to go to my room | n | 143 | 175 | 200 | 225 | 240 |
|  | % | 14.5 | 17.8 | 20.3 | 22.9 | 24.4 |
| **Patient’s Perception of Waiting times and delays** | | | | | | |
| A5. I felt that my scheduled tests and procedures were performed on time | n | 144 | 163 | 218 | 220 | 238 |
|  | % | 14.6 | 16.6 | 22.2 | 22.4 | 24.2 |
| A6. I was kept informed about the results of tests and treatments on time | n | 166 | 116 | 211 | 213 | 277 |
|  | % | 16.9 | 11.8 | 21.5 | 21.7 | 28.2 |
| A7. My discharge was handled in a timely manner | n | 162 | 188 | 207 | 222 | 204 |
|  | % | 16.5 | 19.1 | 21.1 | 22.6 | 20.8 |
| A8. The medical procedures were done correctly the first time | n | 149 | 156 | 237 | 216 | 225 |
|  | % | 15.2 | 15.9 | 24.1 | 22 | 22.9 |
| A9. When I needed help getting, I got help in time | n | 147 | 195 | 222 | 233 | 186 |
|  | % | 15.0 | 19.8 | 22.6 | 23.7 | 18.9 |
| **Patient’s Perception of Availability and Accessibility** | | | | | | |
| A10. My physicians spent an appropriate amount of time with me | n | 143 | 119 | 265 | 218 | 238 |
|  | % | 14.5 | 12.1 | 27.0 | 22.2 | 24.2 |
| A11. The doctor who treated me was available all the time | n | 142 | 168 | 229 | 220 | 224 |
|  | % | 14.4 | 17.1 | 23.3 | 22.4 | 22.8 |
| A12. The doctor who treated me was available during holidays or weekends | n | 149 | 173 | 168 | 238 | 255 |
|  | % | 15.2 | 17.6 | 17.1 | 24.2 | 25.9 |
| A13. There was a specialist present in all shifts, regardless of shift time | n | 143 | 159 | 231 | 228 | 222 |
|  | % | 14.5 | 16.2 | 23.5 | 23.2 | 22.6 |
| A14. There was one particular doctor in charge of my care | n | 138 | 160 | 229 | 217 | 239 |
|  | % | 14.0 | 16.3 | 23.3 | 22.1 | 24.3 |
| A15. Then my family needed to confer with doctors, the doctors were available | n | 140 | 109 | 238 | 225 | 271 |
|  | % | 14.2 | 11.1 | 24.2 | 22.9 | 27.6 |
| **Patient’s Perception of Transition to Home** | | | | | | |
| TR1. I received written instructions when I was sent home | n | 140 | 152 | 260 | 224 | 207 |
|  | % | 14.2 | 15.5 | 26.4 | 22.8 | 21.1 |
| TR2. Someone explained the purpose of the medicine I had to take at home in a way I could understand | n | 170 | 136 | 231 | 226 | 220 |
|  | % | 17.3 | 13.8 | 23.5 | 23.0 | 22.4 |
| TR3. Someone on the hospital staff told Tell me what danger signals to watch for after I go home | n | 150 | 134 | 272 | 176 | 251 |
|  | % | 15.3 | 13.6 | 27.7 | 17.9 | 25.5 |
| TR4. Someone told me about the side effects to watch once I am at home | n | 150 | 140 | 275 | 181 | 237 |
|  | % | 15.3 | 14.2 | 28.0 | 18.4 | 24.1 |
| TR5. Someone told me when I could resume my usual activities (go back to work or drive) | n | 128 | 128 | 290 | 206 | 231 |
|  | % | 13.0 | 13.0 | 29.5 | 21.0 | 23.5 |
| TR6. The doctors gave my family all the information they needed to help me recover | n | 150 | 177 | 241 | 215 | 200 |
|  | % | 15.3 | 18.0 | 24.5 | 21.9 | 20.3 |
| TR7. The nurses gave my family all the information they needed to help me recover | n | 134 | 171 | 256 | 178 | 244 |
|  | % | 13.6 | 17.4 | 26 | 18.1 | 24.8 |
| **Patient’s perception of Management rules and regulations** | | | | | | |
| O1- I feel that the hospital conducts more medical tests than necessary | n | 133 | 154 | 245 | 245 | 206 |
|  | % | 13.5 | 15.7 | 24.9 | 24.9 | 21.0 |
| O2- I feel that the hospital rules were strictly maintained. | n | 151 | 162 | 262 | 154 | 254 |
|  | % | 15.4 | 16.5 | 26.7 | 15.7 | 25.8 |
| O3- I noticed a lack of coordination between units or sections in the hospital | n | 138 | 172 | 240 | 207 | 226 |
|  | % | 14.0 | 17.5 | 24.4 | 21.1 | 23.0 |
| O4- The administrative procedures were done correctly the first time | n | 128 | 156 | 240 | 213 | 246 |
|  | % | 13 | 15.9 | 24.4 | 21.7 | 25.0 |
| O5- The admission process was quite organized | n | 152 | 146 | 236 | 202 | 247 |
|  | % | 15.5 | 14.9 | 24.0 | 20.5 | 25.1 |
| O6- The working hours for the cafeteria were convenient | n | 145 | 177 | 237 | 209 | 215 |
|  | % | 14.8 | 18.0 | 24.1 | 21.3 | 21.9 |
| O7- Visiting rules and regulations were always enforced by the hospital staff | n | 134 | 153 | 248 | 236 | 212 |
|  | % | 13.6 | 15.6 | 25.2 | 24.0 | 21.6 |
| O8- Visiting rules and regulations were enforced at all units (i.e., ICU, Emergency, etc.) | n | 162 | 193 | 174 | 242 | 212 |
|  | % | 16.5 | 19.6 | 17.7 | 24.6 | 21.6 |
| **Patient’s perception of Payment matters** | | | | | | |
| O9- When I had to pay for the treatment, I got as much help as I wanted from someone on the hospital staff in figuring out how to pay my hospital bills | n | 156 | 162 | 215 | 242 | 208 |
|  | % | 15.9 | 16.5 | 21.9 | 24.6 | 21.2 |
| O10- When I had to pay for the treatment, the procedures were simple | n | 144 | 144 | 238 | 235 | 222 |
|  | % | 14.6 | 14.6 | 24.2 | 23.9 | 22.6 |
| O11- When I had to pay for the treatment, the overall cost was reasonable | n | 139 | 131 | 269 | 230 | 214 |
|  | % | 14.1 | 13.3 | 27.4 | 23.4 | 21.8 |
| **Patient’s perception of Outcome and overall assessment (satisfaction And Loyalty):** | | | | | | |
| S1. I am satisfied with the treatment and outcome of care | n | 149 | 133 | 250 | 178 | 273 |
|  | % | 15.2 | 13.5 | 25.4 | 18.1 | 27.8 |
| S2. Compared to other hospitals, this hospital is considered to be a good place for care | n | 163 | 173 | 213 | 229 | 205 |
|  | % | 16.6 | 17.6 | 21.7 | 23.3 | 20.9 |
| S3. I am satisfied with the care provided by this hospital | n | 176 | 172 | 207 | 183 | 245 |
|  | % | 17.9 | 17.5 | 21.1 | 43 | 24.9 |
| S4. I received courteous and professional care while in the hospital | n | 149 | 181 | 237 | 139 | 277 |
|  | % | 15.2 | 18.4 | 24.1 | 14.1 | 28.2 |
| S5. Overall, I was satisfied with the care I received at this hospital | n | 149 | 149 | 254 | 204 | 227 |
|  | % | 15.2 | 15.2 | 25.8 | 20.8 | 23.1 |
| L1. I would be willing to return to this hospital in the future if needed | n | 154 | 144 | 234 | 172 | 279 |
|  | % | 15.7 | 14.6 | 23.8 | 17.5 | 28.4 |
| L2. I feel comfortable recommending the hospital to my friend | n | 158 | 140 | 221 | 195 | 269 |
|  | % | 16.1 | 14.2 | 22.5 | 19.8 | 27.4 |
| L3. I would recommend the hospital to my family | n | 167 | 141 | 246 | 162 | 267 |
|  | % | 17.0 | 14.3 | 25.0 | 16.5 | 27.2 |

Table S3: The reliability of the study's constructs using Cronbach’s Alpha

|  | Mean | Minimum | Maximum | SD | n of Items | Cronbach’s alpha |
| --- | --- | --- | --- | --- | --- | --- |
| Tangible | 3.14 | 3.017 | 3.268 | 0.084 | 13 | 0.927 |
| Empathy | 3.19 | 3.103 | 3.265 | 0.063 | 7 | 0.875 |
| Responsiveness | 3.08 | 2.990 | 3.201 | 0.063 | 8 | 0.908 |
| Competency | 3.18 | 3.090 | 3.268 | 0.045 | 11 | 0.916 |
| Information | 3.19 | 3.135 | 3.302 | 0.063 | 7 | 0.877 |
| Availability | 3.26 | 3.113 | 3.435 | 0.084 | 15 | 0.92 |
| Transition | 3.24 | 3.159 | 3.315 | 0.045 | 7 | 0.884 |
| Organization | 3.23 | 3.135 | 3.284 | 0.045 | 11 | 0.915 |
| Patient Satisfaction | 3.22 | 3.154 | 3.333 | 0.071 | 5 | 0.849 |
| Patient Loyalty | 3.26 | 3.227 | 3.293 | 0.032 | 3 | 0.769 |
| Healthcare quality | 3.19 | 2.990 | 3.435 | 0.084 | 79 | 0.937 |

Table S4: Regression satisfaction model fit

| **Model** | **R** | **R²** | **Adjusted R²** | **Std. Error of Estimate** | **Durbin–Watson** |
| --- | --- | --- | --- | --- | --- |
| 1 | 0.759 | 0.577 | 0.573 | 0.696 | 1.028 |

Table S5: Residuals Statistics for satisfaction model

| **Statistic** | **Minimum** | **Maximum** | **Mean** | **SD** | **n** |
| --- | --- | --- | --- | --- | --- |
| Predicted Value | 0.883 | 5.788 | 3.205 | 0.809 | 983 |
| Residual | -2.932 | 1.837 | 0.000 | 0.693 | 983 |
| Std. Predicted | -2.869 | 3.192 | 0.000 | 1.000 | 983 |
| Std. Residual | -4.212 | 2.639 | 0.000 | 0.996 | 983 |

Table S6. Residual Statistics for Loyalty Model

| **Statistic** | **Minimum** | **Maximum** | **Mean** | **Std. Deviation** | **n** |
| --- | --- | --- | --- | --- | --- |
| Predicted Value | 0.9384 | 5.4375 | 3.2631 | 0.92018 | 983 |
| Residual | -2.6977 | 2.1387 | 0.0000 | 0.67894 | 983 |
| Standardized Predicted | -2.526 | 2.363 | 0.000 | 1.000 | 983 |
| Standardized Residual | -3.955 | 3.136 | 0.000 | 0.995 | 983 |
