## Supplementary figures and images for "Perceived Healthcare Quality, Patient Satisfaction, and Loyalty in Egypt’s Health Insurance Organization: A Multivariate Analysis"

### Suppl figures

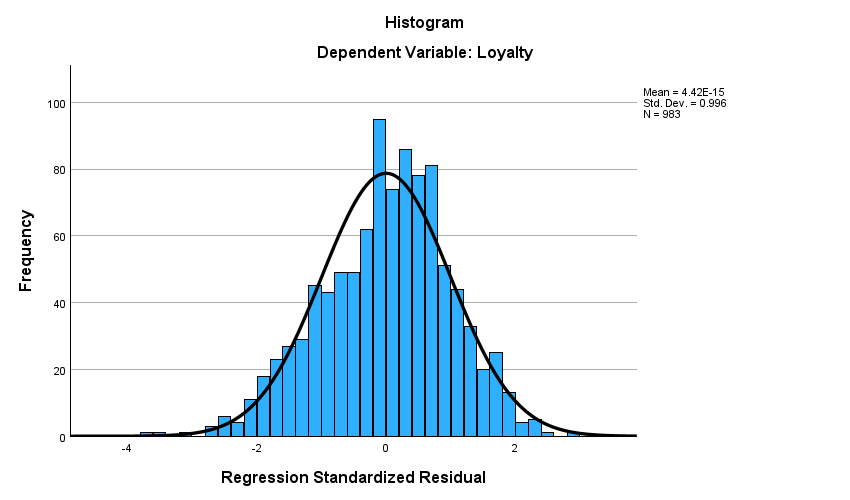

Figure S1

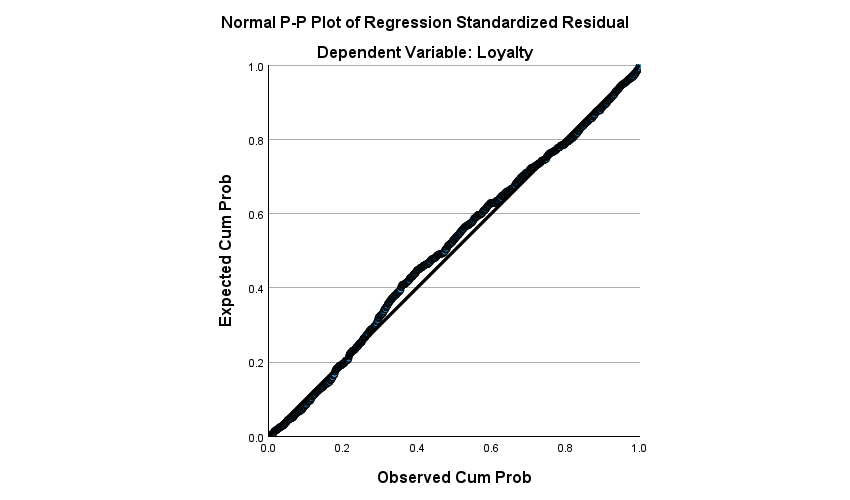

Figure S2

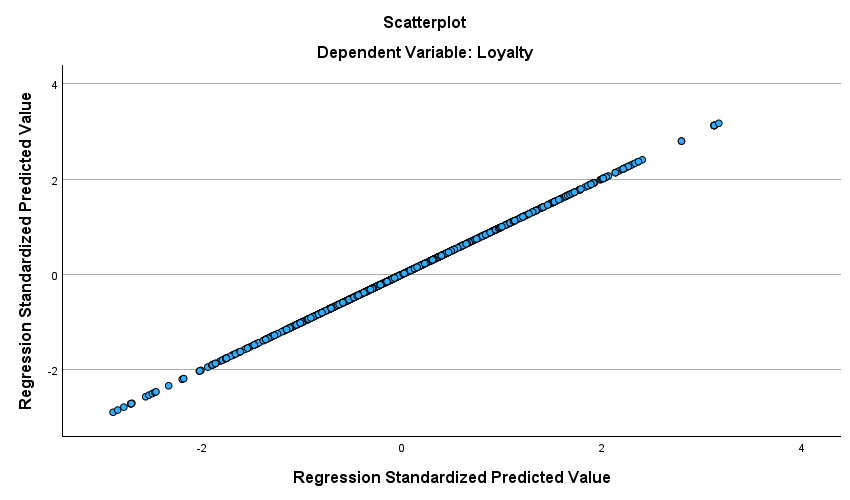

Figure S3
